# Association between health-related hope and adherence to prescribed treatment in CKD patients: multicenter cross-sectional study

**DOI:** 10.1101/2020.07.13.20130104

**Authors:** Noriaki Kurita, Takafumi Wakita, Yoshitaka Ishibashi, Shino Fujimoto, Masahiko Yazawa, Tomo Suzuki, Kenichiro Koitabashi, Mai Yanagi, Hiroo Kawarazaki, Joseph Green, Shunichi Fukuhara, Yugo Shibagaki

**Author notes:** **Address correspondence to:** Noriaki Kurita, MD, PhD, FACP, Department of Clinical Epidemiology, Graduate School of Medicine, Fukushima Medical University, 1 Hikarigaoka, Fukushima City, Fukushima 960-1295, Japan, **E-mail:**.

## Abstract

**Background:** In chronic kidney disease (CKD), patients’ adherence to prescriptions for diet and for medications might depend on the degree to which they have hope that they will enjoy life, and that hope could vary with the stage of CKD. The aims of this study were to quantify both the association of CKD stage with health-related hope (HR-Hope), and the association of that hope with psychological and physiological manifestations of adherence.

**Methods:** This was a cross-sectional study involving 461 adult CKD patients, some of whom were receiving dialysis. The main exposure was HR-Hope, measured using a recently-developed 18-item scale. The outcomes were perceived burden of fluid restriction and of diet restriction, measured using the KDQOL, and physiological manifestations of adherence (systolic and diastolic blood pressure [BP], and serum phosphorus and potassium levels).

General linear models and generalized ordered logit models were fit.

**Results:** Participants at non-dialysis stage 4 and those at stage 5 had lower HR-Hope scores than did those at stage 2 or 3 (combined). Those at non-dialysis stage 5 had the lowest scores. HR-Hope scores of participants at stage 5D were similar to those of participants at stage 4, but they were lower than the scores of participants at stage 2 or 3 (combined). Higher HR-Hope scores were associated with lower perceived burdens of fluid restriction and of diet restriction (adjusted ORs per ten-point difference were 0.82 and 0.84, respectively). Higher HR-Hope scores were associated with lower systolic BP (adjusted mean difference in systolic BP per ten-point difference in HR-Hope scores was -1.87 mmHg). In contrast, HR-Hope scores were not associated with diastolic BP, serum phosphorus levels, or serum potassium levels.

**Conclusions:** Among CKD patients, HR-Hope is associated with disease stage, with psychological burden, and with some physiological manifestations of adherence.

## Background

In patients with chronic kidney disease (CKD), mortality and cardiovascular morbidity can be reduced. Specifically, they can be reduced if increases in blood pressure, retention of fluid, and excessive accumulation of salt, potassium, and phosphorus are prevented. Those goals can be achieved at least to some extent by adherence – both adherence to prescriptions for medication and adherence to prescribed restrictions on the intake of fluids and solid foods. (1, 2) Unfortunately, non-adherence appears to be quite common. (3) Results of a qualitative study indicate that both non-adherence to prescribed medication intake and non-adherence to prescribed fluid and dietary intake may be due in part to lack of motivation. (2) Research on other chronic diseases identified at least one important source of motivation for adherence: patients’ hope. (4, 5)

Hope can be regarded as a goal-oriented way of thinking that enables people both to find routes toward their goals and also to sustain their motivation to pursue those goals. (6, 7) More than hope among healthy people, hope among patients with chronic illnesses is concerned with health. (8) For patients with CKD, it is possible that adherence to prescriptions for diet and medications can help them achieve their hoped-for goals. Thus, CKD patients with greater health-related hope may better cope with the burdens of adherence to those prescriptions, and as a result they might be better able to maintain relatively good health. Despite this theoretical importance of hope in CKD patients, it has been studied only rarely. (9)

One analytical cross-sectional study of patients at CKD stage 5D showed that high scores on Snyder’s hope scale were associated with a lower burden of kidney disease as measured using the Kidney Disease Quality of Life (KDQOL) instrument. (9) That study included only CKD patients who were undergoing dialysis. Their symptoms were not correlated with scores on Snyder’s hope scale (which measures hope in general), but other likely correlates of hope, such as the severity of CKD and physiological manifestations of adherence to prescribed medications and diet, were not examined. (9) Patients’ experiences of CKD vary with the stage of the disease: Patients who have some hope for enjoying life have a tendency to adjust to their condition, or to control it. (10) In this context, studies quantifying health-related hope, its relationships with patients’ experiences at different stages of CKD, and its associations with physiological manifestations of adherence to prescribed medications and diet are likely to be clinically relevant. Specifically, such studies might help patients and clinicians develop interventions to increase hope and thereby promote adherence to those prescriptions.

To quantify the association of CKD stage with health-related hope, and the association of that hope with psychological and physiological manifestations of adherence to prescribed medications and diet, we used a recently developed scale to measure health-related hope (HR-Hope) (11) and we analyzed cross-sectional data from a prospective cohort study: the HOpe Trajectory and Disease Outcome Consortium (HOTDOC) for Japanese patients with CKD.

## Methods

This was a multicenter cross-sectional study. The protocol was approved by the ethics review boards of the local health authorities.

### Target population and settings

All of the participants were adults with CKD who were being treated by nephrologists. Some did not require dialysis, while others had CKD stage 5D and were receiving hemodialysis or peritoneal dialysis. Those with advanced cancer who were likely to die within one year were not included. Also not included were those with a psychiatric condition that could impair their ability to understand or respond to verbal or written instructions (e.g. advanced dementia, schizophrenia, intellectual disability). The settings were one university hospital and four general hospitals. A total of 461 patients were registered between February 2016 and September 2017.

### Study procedures

Patients were asked to respond to the questionnaire after they gave informed consent.

The questionnaire included the HR-Hope scale, and selected items from the Japanese version of the KDQOL instrument.

### Health-related hope scale (HR-Hope)

The HR-Hope scale measures hope among people with chronic diseases (Table S1 and S2). (11) The items use a 4-point Likert-type scale: I don’t feel that way at all, I feel that way a little, I feel that way somewhat, I strongly feel that way. The respondents are asked to “Please answer the questions below while keeping in mind how you feel about your future health prospects”. Internal consistency reliability (coefficient alpha) was 0.93. Scores on the HR-Hope scale were moderately correlated with scores on both domains of Snyder’s hope scale, (7, 12) and with scores on the 8 domains of the Medical Outcomes Study SF-36.(13) In addition, the HR-Hope scale was more sensitive to change in socio-clinical status than was Snyder’s hope scale. For this study, the mean score of all 18 items was computed. For patients without family, the 2 items related to family were not applicable, thus the scale score was computed as the mean of the remaining 16 items. Next, the mean score was transformed to a 0 – 100 score.

### Exposures and outcomes: analytic framework

We examined how HR-Hope was associated with the psychological burden of CKD and with physiological manifestations of adherence that are applicable across all stages of kidney disease (see the conceptual framework in Fig. S1): (1-i) burden of fluid restriction, (1-ii) burden of diet restriction, (2-i) systolic blood pressure (BP), (2-ii) diastolic BP, (3-i) serum phosphorus levels, and (3-ii) serum potassium levels. Abnormalities in serum phosphorus and potassium levels were taken as manifestations of non-adherence to prescribed diet. (14) Because excessive fluid and salt intake, and skipping prescribed doses of anti-hypertensive medications can contribute to BP elevation among CKD patients, abnormally high systolic and diastolic BPs were taken as manifestations of non-adherence to prescriptions for medications, fluid intake, and salt intake. Six categories of CKD were considered: non-dialysis stage 2/3, non-dialysis stage 4, non-dialysis stage 5, dialysis for 1 year or less, dialysis for >1 to 3 years, and dialysis for > 3 years. The perceived burdens of fluid restriction and diet restriction were assessed using items from the “Burden” subscale of the KDQOL.(15, 16) Participants were asked “How much does kidney disease bother you in each of the following areas?” and the two areas considered were fluid restriction and dietary restriction. The response choices were “Not at all bothered”, “Somewhat bothered”, “Moderately bothered”, “Very much bothered”, and “Extremely bothered”. Next, we examined whether HR-Hope was associated with the stage of CKD and with key socio-demographic factors (see the conceptual framework in Fig. S1).

### Covariates

Covariates used in the analyses included socio-demographic characteristics (age, gender, presence of family, and working status as a proxy for socioeconomic status), primary renal disease, comorbidities (diabetes, coronary artery disease, and cerebrovascular disease), performance status (PS), and medication (type of phosphate binder, number of phosphate binders prescribed, prescription of potassium binder, class of antihypertensives prescribed, and total number of classes of antihypertensives). Presence of family was asked with the item “Do you have any family?” and the patients chose yes/no. Working status was asked using an item from KDQOL,(15, 16) in which the patients answered yes/no to “During the past 4 weeks, did you work at a paying job?”. PS was assessed by the attending physician using scores developed by the Eastern Cooperative Oncology Group (Zubrod Scale). (17) Possible scores range from 0 (normal activity) to 4 (bedridden). A score of 2 (Ambulatory and capable of all self-care but unable to carry out any work activities. Up and about more than 50% of waking hours.) or higher was defined as impaired PS.

#### Data collection

Medical data were collected from medical records by trained staff. Data on socio-demographic factors, primary renal disease, dialysis duration (only for patients receiving hemodialysis or peritoneal dialysis), comorbidities, medications, BP, and serum phosphorus and potassium were extracted from medical records written at the time when the participants received the questionnaires during their hospital visits. Among those receiving hemodialysis, the data on BP and on serum phosphorus and potassium were those collected routinely, before the first dialysis session of each week (typically before starting dialysis on a Monday or a Tuesday).

### Statistical analysis

All statistical analyses were done using Stata/SE version 15 (Stata Corp., College Station, TX). Socio-demographic characteristics, comorbidities, laboratory data, and medication data were summarized as means and standard deviations for continuous variables and percentages for categorical variables. The distributions of degree of burdens of fluid or diet restriction by quartile-defined categories of HR-Hope score were graphed. Trends across those categories of HR-Hope score were analyzed using a non-parametric trend test for degree of burdens of fluid or diet restriction. Associations of HR-Hope with the perceived burdens of fluid and diet restriction were analyzed using generalized ordered logit models, (18) with and without adjustment for key socio-demographic variables (adjusted model 1), and for variables in adjusted model 1 together with primary renal disease and comorbidities (adjusted model 2). (18) For these analyses, the responses were collapsed into three categories: “Not at all bothered”, “Somewhat or Moderately bothered”, and “Very much or Extremely bothered”, and odds ratios for increasing likelihood of degree of burden (at least “Somewhat or Moderately bothered” or at least “Very much or Extremely bothered”) were estimated.

The analytic models of the associations of HR-Hope with systolic and diastolic BP were general linear models, with and without adjustment for key socio-demographic variables (adjusted model 1), and for the variables in adjusted model 1, primary renal disease, comorbidities, and the number of classes of prescribed antihypertensives (adjusted model 2). The analytic models of the association of HR-Hope with serum phosphorus were general linear models, with and without adjustment for key socio-demographic variables (adjusted model 1), and for the variables in adjusted model 1, primary renal disease, comorbidities, and the number of prescribed phosphate binders (adjusted model 2). The analytic models of the association of HR-Hope with serum potassium were general linear models, with and without adjustment for key socio-demographic variables (adjusted model 1), and for the variables in adjusted model 1, primary renal disease, comorbidities, and prescription of potassium binders (adjusted model 2). The analytic models of the association of categories of CKD with HR-Hope were general linear models, with and without adjustment for key socio-demographic variables (adjusted model 1), and for the variables in adjusted model 1 together with primary renal disease and comorbidities (adjusted model 2). Within each analysis, all problems of missing data on covariates were addressed by multiple imputation. (19) Five imputations were done using chained equations, with the assumption that the data were missing at random. P values < 0.05 were taken as indicators of statistical significance.

## Results

### Participants

Characteristics of the 461 participants are shown in Table 1. About 10.2% of the participants had impaired PS. Among the dialysis patients, about one third were receiving peritoneal dialysis (32%). Compared with the non-dialysis patients, the dialysis patients were younger, and they were more likely to have diabetic nephropathy, impaired PS, and higher systolic BP and phosphorus levels. HR-Hope scores were normally distributed (Figure 1).

**Table 1.** Characteristics of analysis population.

|  | Non-dialysis<br>Stage 2/3<br>n = 59 | Non-dialysis<br>Stage 4<br>n = 55 | Non-dialysis<br>Stage 5<br>n = 19 | Dialysis<br>0 to 1 yr<br>n = 66 | Dialysis<br>>1 to 3yr<br>n = 72 | Dialysis<br>>3 yr<br>n = 190 |
| --- | --- | --- | --- | --- | --- | --- |
| **Dialysis modality, %<sup>a</sup>** |  |  |  |  |  |  |
| Hemodialysis |  |  |  | 71.2% | 52.8% | 72.6% |
| Peritoneal dialysis |  |  |  | 28.8% | 47.2% | 27.4% |
| **eGFR, mL/min/1.73 m<sup>2</sup>** | 46.1<br>(12.3) | 22.4<br>(4.3) | 11.5<br>(2.8) | 6.9<br>(2.7) | 5.0<br>(2.1) | 4.0<br>(1.5) |
| **Age, yr** | 70.7<br>(12.7) | 74.5<br>(12.5) | 73.9<br>(9.2) | 66.8<br>(15.5) | 64.9<br>(15.5) | 64.3<br>(12.7) |
| **Women, %** | 33.9% | 30.9% | 36.8% | 27.3% | 22.2% | 36.3% |
| **Renal disease, %** |  |  |  |  |  |  |
| Diabetic nephropathy | 5.1% | 21.8% | 15.8% | 30.3% | 43.1% | 26.8% |
| Glomerulonephritis | 13.6% | 12.7% | 21.1% | 25.8% | 23.6% | 34.2% |
| Nephrosclerosis | 37.3% | 32.7% | 31.6% | 27.3% | 15.3% | 6.8% |
| Polycystic kidney | 0.0% | 0.0% | 5.3% | 7.6% | 1.4% | 7.9% |
| Others/Unknown | 44.1% | 32.7% | 26.3% | 9.1% | 16.7% | 24.2% |
| **Impaired performance status, %** | 1.7% | 5.5% | 5.3% | 13.6% | 6.9% | 14.7% |
| **Diabetes, %** | 18.6% | 45.5% | 31.6% | 43.9% | 50.0% | 33.2% |
| **Cerebrovascular disease, %** | 6.8% | 16.4% | 15.8% | 16.7% | 16.7% | 12.1% |
| **Coronary artery disease, %** | 3.4% | 23.6% | 15.8% | 16.7% | 15.3% | 16.8% |
| **SBP, mmHg** | 130<br>(17) | 133<br>(17) | 142<br>(20) | 139<br>(22) | 143<br>(24) | 145<br>(28) |
| *Missing* | 12 | 11 | 5 |  | 1 |  |
| **DBP, mmHg** | 79 | 69 | 72 | 75 | 75 | 79 |
|  | (12) | (14) | (12) | (15) | (15) | (17) |
| *Missing* | 23 | 21 | 5 |  | 1 |  |
| **Phosphorus, mg/dL** | 3.2 | 3.6 | 4.0 | 5.3 | 5.4 | 5.5 |
|  | (0.4) | (0.5) | (0.7) | (1.5) | (1.3) | (1.4) |
| *Missing* | 31 | 5 |  |  |  | 1 |
| **Potassium, mEq/L** | 4.4 | 4.8 | 4.7 | 4.4 | 4.4 | 4.7 |
|  | (0.3) | (0.6) | (0.5) | (0.7) | (0.7) | (0.8) |
| *Missing* | 3 |  |  |  |  | 2 |
| **N of categories of antihypertensive drugs, %** |  |  |  |  |  |  |
| None | 39.0% | 14.6% | 15.8% | 21.2% | 23.6% | 38.4% |
| 1 | 33.9% | 29.1% | 26.3% | 28.8% | 23.6% | 25.8% |
| 2 | 20.3% | 32.7% | 52.6% | 31.8% | 34.7% | 21.1% |
| 3 or more | 6.8% | 23.6% | 5.3% | 18.2% | 18.1% | 14.7% |
| **N of phosphate binder, %** |  |  |  |  |  |  |
| None | 100.0% | 100.0% | 89.5% | 68.2% | 30.6% | 16.3% |
| 1 to 5 | 0.0% | 0.0% | 10.5% | 18.2% | 25.0% | 19.5% |
| 6 to 10 | 0.0% | 0.0% | 0.0% | 13.6% | 27.8% | 41.6% |
| 11 to 15 | 0.0% | 0.0% | 0.0% | 0.0% | 9.7% | 15.8% |
| 16 or over | 0.0% | 0.0% | 0.0% | 0.0% | 6.9% | 6.8% |
| **Prescription for potassium binder, %** | 1.7% | 16.4% | 5.3% | 3.0% | 13.9% | 23.7% |

For continuous variable, data were summarized as mean and standard deviation in parenthesis.
<sup>a</sup>Calculated only among dialysis patients.
eGFR: estimated glomerular filtration rate; SBP: systolic blood pressure; diastolic blood pressure

**Figure 1.**
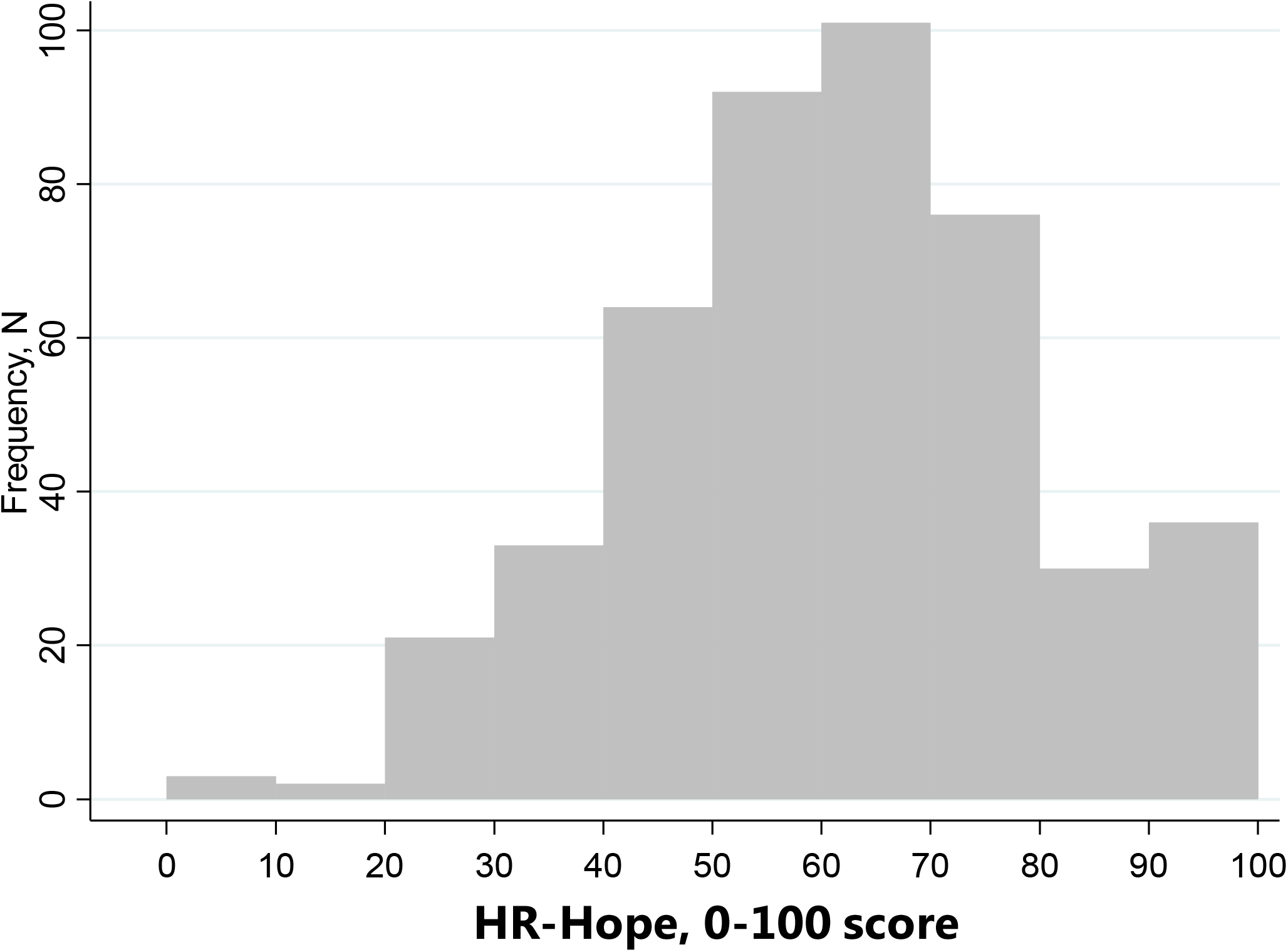
Distribution of HR-Hope scores

### Associations between HR-Hope and burdens

Figure 2 shows the distribution of perceived burden of fluid and dietary restrictions, by quartile-defined categories of HR-Hope scores. Both burdens were lower among participants with higher HR-Hope scores (lower burdens of fluid and dietary restrictions were associated with higher HR-Hope scores: *p* < 0.001 and *p* = 0.001, respectively). Those associations were unchanged after adjustment for likely confounders (adjusted odds ratio [OR] per ten-point difference 0.82, 95% confidence interval [CI] 0.73 to 0.92 for fluid restriction, and 0.84, 95%CI 0.76 to 0.94 for dietary restriction, Table 2). Older participants perceived fluid restriction as being less burdensome than did younger participants (adjusted OR per ten-year difference 0.79, 95%CI 0.66 to 0.94). Employed participants perceived dietary restriction as being more burdensome than did non-employed participants (adjusted OR 1.88, 95%CI 1.20 to 2.94). Participants who had family did not perceive the fluid and dietary restrictions as being more burdensome than participants who did not have family.

**Figure 2.**
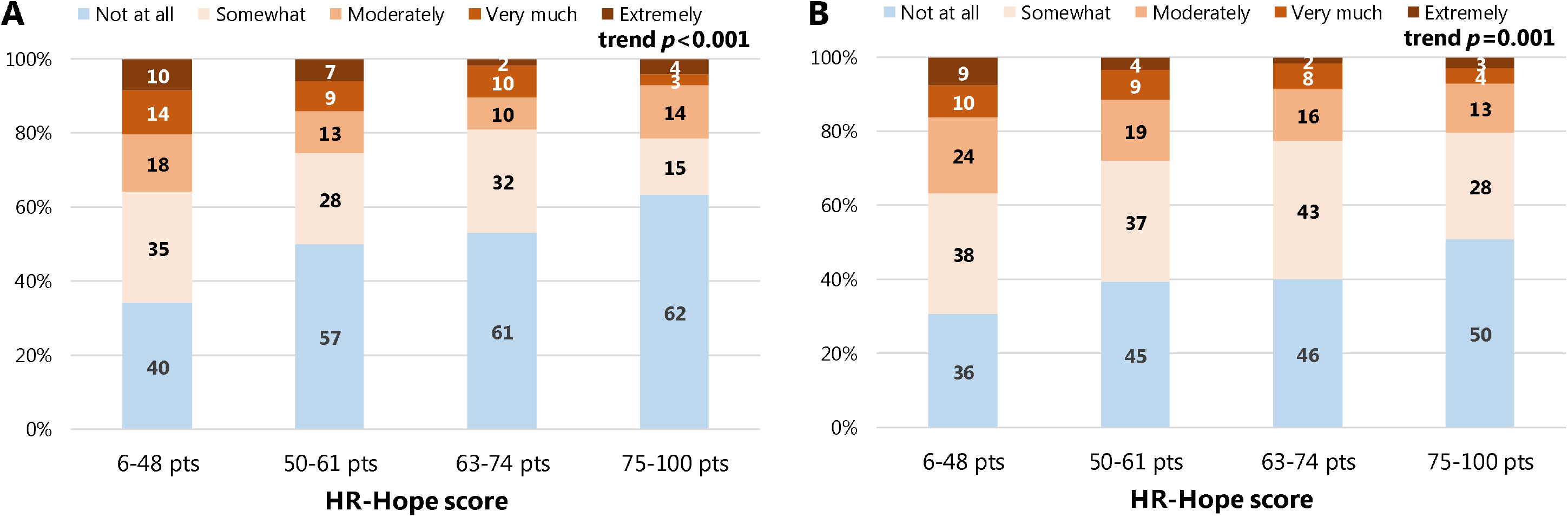
The distributions of degree of burdens of fluid or diet restriction by quartile-defined categories of HR-Hope score. Figure 2A shows distributions of perceived burden of fluid restriction. Figure 2B shows distributions of perceived burden of diet restriction. Vertical axes indicate proportions of responses. Numbers in each bar graph indicate absolute numbers of responses (n = 444). HR-Hope: health-related hope.

**Table 2.** Associations between HR-Hope, key demographic factors, and degree of burdens of fluid or diet restriction.^a^.

|  | Fluid restriction (n = 444), OR (95% CI) |  |  | Diet restriction (n = 444), OR (95% CI) |  |  |
| --- | --- | --- | --- | --- | --- | --- |
|  | Unadjusted | Adjusted 1<sup>b</sup> | Adjusted 2<sup>c</sup> | Unadjusted | Adjusted 1<sup>b</sup> | Adjusted 2<sup>c</sup> |
| **HR-Hope** |  |  |  |  |  |  |
| *per 10 points* | **0.79**<br>(0.71 – 0.87) | **0.83**<br>(0.74 – 0.93) | **0.82**<br>(0.73 – 0.92) | **0.84**<br>(0.76 – 0.93) | **0.85**<br>(0.77 – 0.95) | **0.84**<br>(0.76 – 0.94) |
| *per 1 SD* | **0.65**<br>(0.54 – 0.78) | **0.71**<br>(0.58 – 0.87) | **0.69**<br>(0.56 – 0.85) | **0.72**<br>(0.60 – 0.87) | **0.74**<br>(0.61 – 0.90) | **0.73**<br>(0.60 – 0.89) |
| **Age, yr** |  | **0.80** | **0.79** |  | 0.91 | 0.88 |
| *per 10 yr* | - | (0.68 – 0.95) | (0.66 – 0.94) | - | (0.78 – 1.06) | (0.75 – 1.03) |
| **Presence of family, yes** |  | 0.96 | 1.02 |  | 1.12 | 1.16 |
|  | - | (0.51 – 1.80) | (0.53 – 1.97) | - | (0.62 – 2.02) | (0.63 – 2.15) |
| **Working, yes** |  | 1.11 | 1.13 |  | **1.80** | **1.88** |
|  | - | (0.69 – 1.79) | (0.69 – 1.85) | - | (1.16 – 2.79) | (1.20 – 2.94) |

<sup>a</sup>Generalized ordered logit models were used to estimate odds ratios for increasing degree of burdens.
<sup>b</sup>Adjusted for age, gender, stage of renal disease, performance status, presence of family, and working status.
<sup>c</sup>Adjusted for the covariates listed in footnote b, and also for primary renal disease, diabetes, coronary artery disease, and cerebrovascular disease.
HR-Hope: health-related hope; OR: odds ratio; 95% CI: 95% Confidence interval.

### Associations between HR-Hope and blood pressure

Participants with higher HR-Hope scores had lower systolic BP (mean difference per ten-point difference -1.9 mmHg, 95%CI -3.1 to -0.7) (Table S3, Fig. 3). In contrast, participants with higher HR-Hope scores had lower diastolic BP in the unadjusted analysis (mean difference per ten-point difference -0.9 mmHg, 95%CI -1.8 to -0.1), but not after adjustment (mean difference per ten-point difference -0.6 mmHg, 95%CI -1.4 to 0.2).

**Figure 3.**
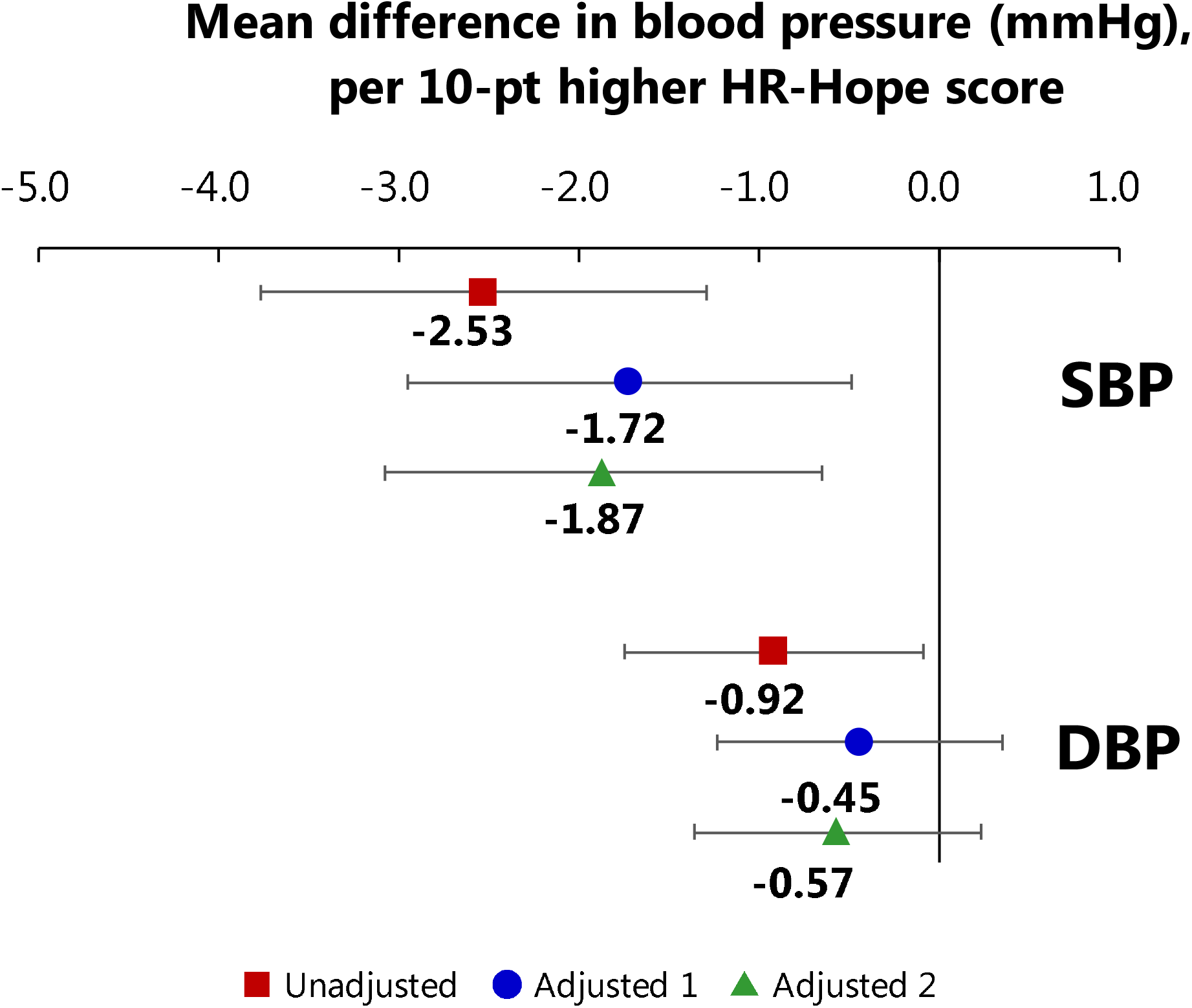
Associations between HR-Hope and blood pressure. Red squares: unadjusted models. Blue circles: models adjusted for age, sex, performance status, presence of family, working status, and number of categories of prescribed antihypertensives (adjusted model 1). Green triangles: models adjusted for all of the variables listed above, and also for primary renal disease, diabetes, coronary artery disease, and cerebrovascular disease (adjusted model 2). Mean differences estimated via general linear models (n = 429 for systolic BP and n = 408 for diastolic BP). Error bars indicate 95% confidence intervals. HR-Hope: health-related hope; SBP: systolic blood pressure, DBP: diastolic blood pressure

Associations between HR-Hope and levels of phosphorus and potassium in serum HR-Hope scores were not associated with the level of phosphorus or potassium in serum (phosphorus: Table S4, Fig. 4A, potassium: Table S5, Fig. 4B).

**Figure 4.**
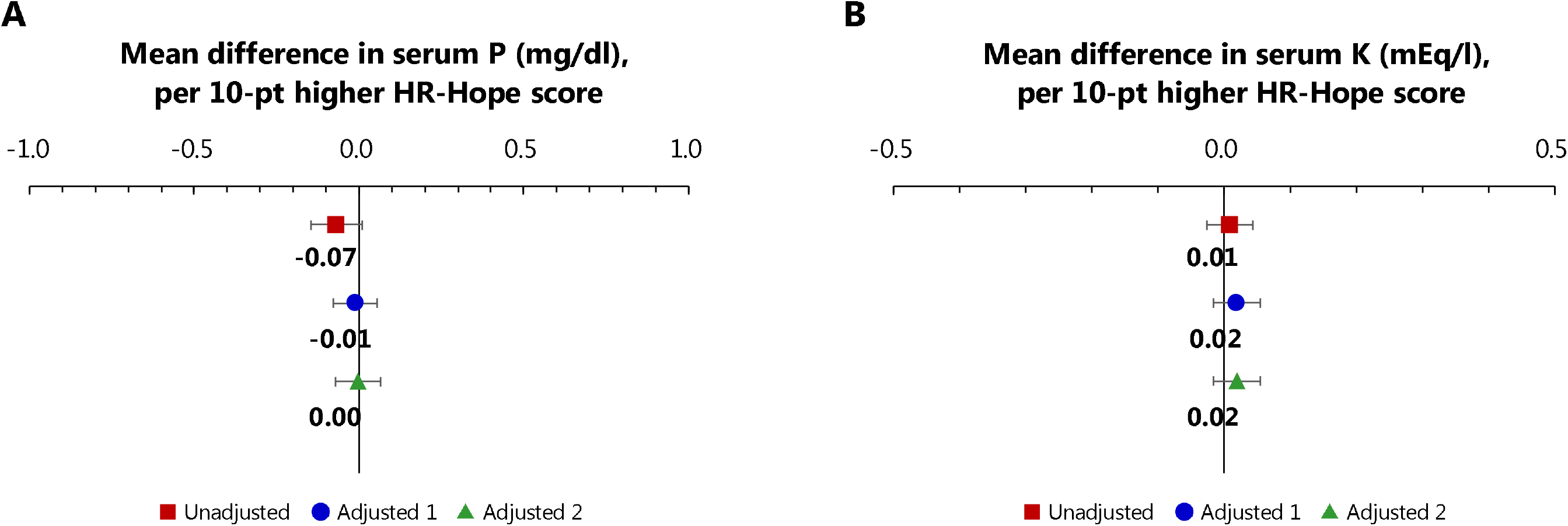
Associations between HR-Hope and serum phosphorus or potassium levels. Figure 4A: Association between HR-Hope and serum phosphorus. Red squares: unadjusted models. Blue circles: models adjusted for age, sex, performance status, presence of family, working status, and number of categories of prescribed phosphate binders (adjusted model 1). Green triangles: models adjusted for all of the variables listed above, and also for primary renal disease, diabetes, coronary artery disease, and cerebrovascular disease (adjusted model 2). Figure 4B: Association between HR-Hope and serum potassium. Red squares: unadjusted models. Blue circles: models adjusted for age, sex, performance status, presence of family, working status, and number of categories of prescribed potassium binders (adjusted model 1). Green triangles: models adjusted for all of the variables listed above, and also for primary renal disease, diabetes, coronary artery disease, and cerebrovascular disease (adjusted model 2). Mean differences estimated via general linear models (n = 422 for phosphorus and n = 453 for potassium). Error bars indicate 95% confidence intervals. HR-Hope: health-related hope; P: phosphorus, K: potassium

### Associations between disease stage and HR-Hope

HR-Hope scores were lower in participants at CKD stages 4 and 5 than in participants at CKD stages 2 and 3 (Table 3, Fig. 5). After adjustment for likely confounders, being at a higher stage of CKD was associated with having a lower HR-Hope score (Fig. 5; mean difference -7.0 points, 95%CI -13.7 to -0.3 for stage 4, and -16.8 points, 95%CI -26.1 to -7.6, for stage 5). The difference in HR-Hope scores between participants at stage 4 and those at stage 2 or 3 was similar to the difference between participants who did and those who did not have family (mean difference 7.7 points, 95%CI 2.3 – 13.1). The standardized effect sizes (Cohen’s *d)* for the differences between HR-Hope scores among participants at stage 4 and at stage 5 versus those at stages 2 and 3 (combined), were 0.38 for stage 4 and 0.91 for stage 5. (20) Among participants on dialysis, the HR-Hope scores were similar to the scores of those at stage 4, but they were lower than the scores of those at stage 2 or 3. Participants who had been receiving dialysis for 1 year or less had lower HR-Hope scores than did those at CKD stage 2 or 3 (mean difference -7.9 points, 95%CI -14.4 to -1.4). While the HR-Hope scores of participants who had been receiving dialysis for between 1 and 3 years were not lower than the scores of those who were at stage 2 or 3 (mean difference -4.9 points, 95%CI -11.3 to 1.6), participants who had been receiving dialysis for 3 years or longer had lower HR-Hope scores than did those at CKD stage 2 or 3 (mean difference -8.8, 95%CI -14.5 to -3.2). Older participants had higher HR-Hope scores than did younger participants (the mean difference was 2.1 points, 95%CI 0.7 to 3.5). The HR-Hope scores of employed participants were not higher than the scores of non-employed participants.

**Table 3.**
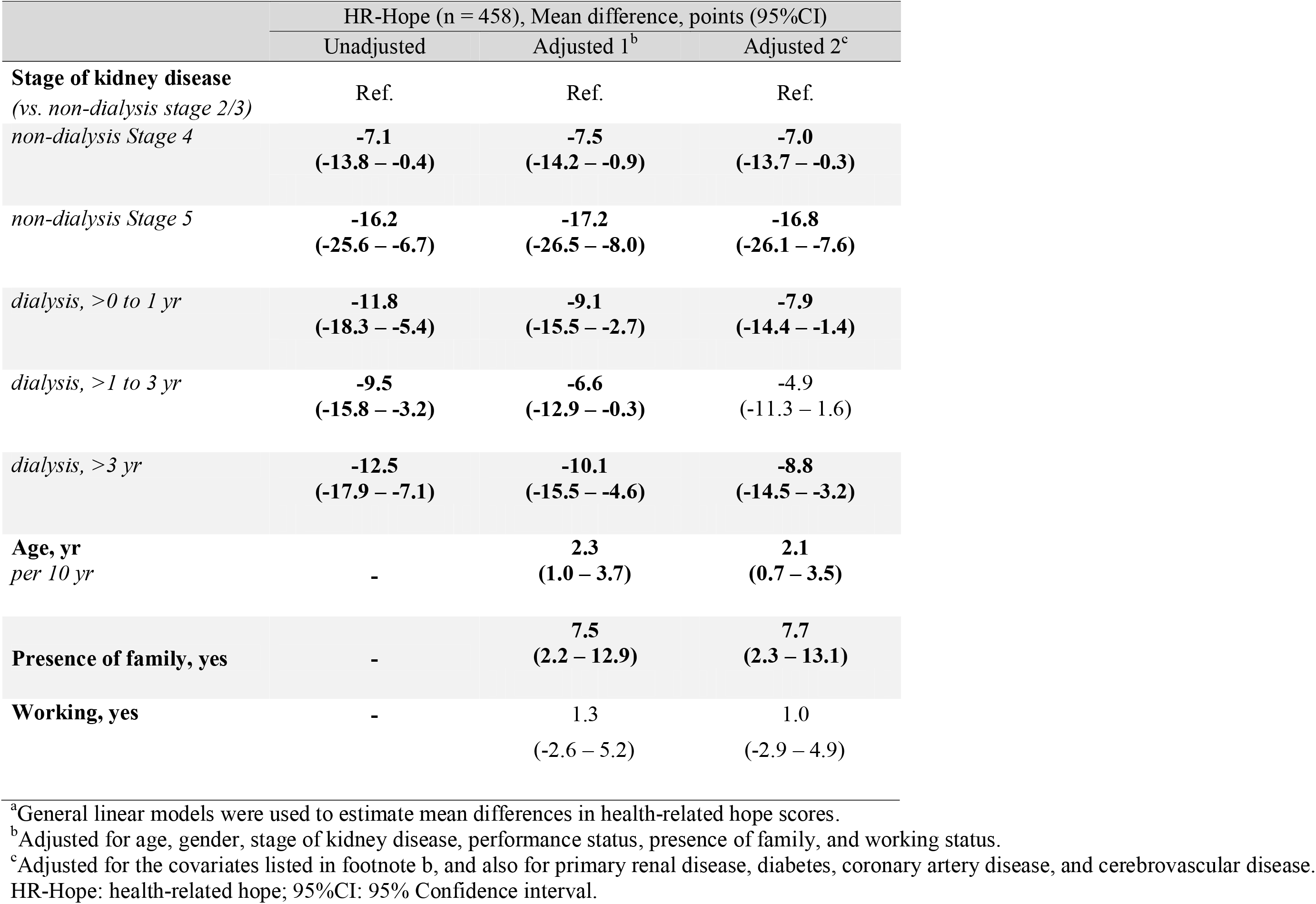
Associations between stage of kidney disease, key demographics, and levels of health-related hope.^a^.

**Figure 5.**
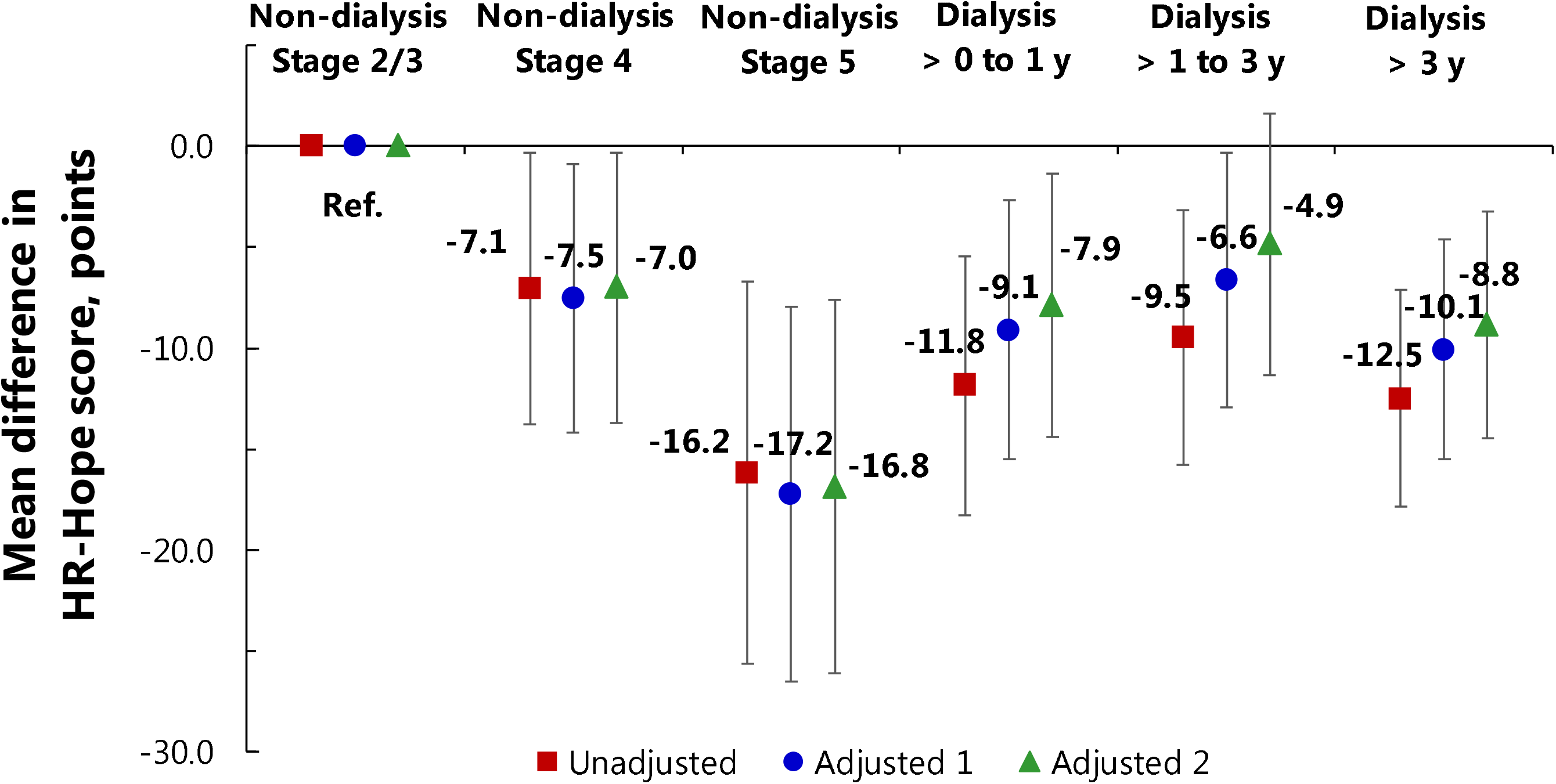
Association between stage of kidney disease and HR-Hope scores. Red squares: unadjusted models. Blue circles: models adjusted for age, sex, performance status, presence of family, and working status (adjusted model 1). Green triangles: models adjusted for all of the variables listed above, and also for primary renal disease, diabetes, coronary artery disease, and cerebrovascular disease (adjusted model 2). Vertical axis indicates mean difference in HR-Hope scores. Error bars indicate 95% confidence intervals. The stages of kidney disease shown here are non-dialysis stages 2 and 3 (combined), which is the reference category, non-dialysis stage 4, non-dialysis stage 5, stage 5D with dialysis duration no longer than 1 year, stage 5D with dialysis duration greater than 1 but no longer than 3 years, and stage 5D with dialysis duration greater than 3 years. Mean differences estimated via general linear models (n = 458). HR-Hope: health-related hope

## Discussion

We found that HR-Hope was greater among those patients who reported that fluid restriction and dietary restriction were less burdensome. In addition, greater HR-Hope was associated with lower systolic BP Finally, HR-Hope was associated with disease stage: hope was lowest among patients at non-dialysis stage 5, and it differed very little among the three groups defined by the duration of dialysis. These findings highlight the importance of integrating consideration of HR-Hope into clinical practice throughout the trajectory of CKD, to achieve better adherence to prescriptions for diet and medication.

These findings may be useful to researchers and clinicians in several ways. First, the finding that lower BP was associated with greater hope (as seen in the adjusted model for systolic BP and in the unadjusted model for diastolic BP) suggests that hopeful patients may more effectively adopt some self-management behaviors, resulting in better BP management. Specifically, that association may have resulted from less salt intake or better adherence to prescriptions for antihypertensive agents, as hopeful patients might be more likely to develop strategies for dealing with their disease and for preventing it from worsening, both of which are measured using the HR-Hope scale (Question 10 and Question 11 in Table S1 and S2). Patients who have specific goals for their health may comprehend self-management behavior as way to achieve their goals and thus they may implement healthy behavior in their daily life.

These notions are partially supported by the results of a previous study, which suggested that dialysis patients who feel benefits from salt restriction actually consume less salt,(21) and those who feel that they can control their health adhere more to prescriptions for medication. (22) Second, hope was not associated with the levels of phosphorus or potassium in serum. Because of the complexity of dietary management (knowledge about amounts of dietary phosphorus and potassium) and simultaneous considerations of family relationships and household income, (2, 23) choosing foods that contain only small amounts of phosphorus and potassium may be difficult. Family relationships can help to support healthy food choices and cooking, but can also interfere with patients’ self-management. (2) Patients who were employed reported feeling a greater burden of dietary restrictions than did those who were not employed. This could reflect employed patients’ difficulty in meal-related socializing. When they gather with work colleagues over lunch or dinner, which is important for maintaining business connections and work-group solidarity, (24) CKD patients may feel obliged to eat as other people do. (25) Third, HR-Hope is potentially modifiable by psychosocial interventions. For example, individualized counseling can be aimed at increasing hope. In one study of hemodialysis patients, a well-organized, regular counseling program helped those patients to frame their hope during their illness and to find ways to reduce anxiety and stress. (26) Fourth, we found that hope was at its minimum among patients at stage 5 CKD, that is, among patients whose disease was so severe that they almost needed dialysis. Also, to the extent that a longitudinal trend can be inferred from data obtained cross-sectionally from patients who had undergone dialysis for different lengths of time (Figure 5), we found that hope was relatively stable over more than 3 years of dialysis therapy. Those two findings suggest that chronicity of disease course might also affect HR-Hope. As renal function deteriorates, HR-Hope might decrease because of anxiety and fear of an imminent lifelong dependence on dialysis, and of burdensome symptoms of uremia such as pulmonary edema, decreased appetite, malaise, and pruritus. While starting dialysis may alleviate symptoms of uremia, patients are forced to adjust their lives to their therapy (i.e. thrice-weekly visits to a dialysis center for those undergoing hemodialysis and every day at home for those undergoing peritoneal dialysis) and to a strict dietary and fluid-intake regimen, and thus they may have less HR-Hope than do patients who are still at stage 2 or 3 of CKD. Our observation that low levels of hope persist during the first 12 months of both hemodialysis and peritoneal dialysis concurs to some degree with previous studies focusing on 12-month change in health-related QOL of dialysis patients. (27, 28) Previous studies have provided conflicting results regarding the impact of being on dialysis for more than 1 year on the mental component of health-related QOL. (29, 30) One cross-sectional study showed that people who had received dialysis for more than 3.5 years had higher mental component summary (MCS) scores than those who had received dialysis for less than 3.5 years. (30) In contrast, in a large longitudinal study people who had received hemodialysis for at least 1 year were no more likely to have improvement in MCS scores than were those who had received hemodialysis for 3 months or less. (29) The reason for the discrepancy between those two sets of results is not clear. However, one reason could be that mental health as measured by the MCS is different from HR-Hope, although mental health and HR-Hope are correlated. (11) Further research is warranted to examine the impact of the duration of dialysis on psychological adjustments, including acceptance and health-related hope.

Several strengths of this study should be noted. First, we showed that HR-Hope is associated with objective manifestations of adherence to prescriptions for diet and medications. In contrast, a previous study of hope in general (not HR-Hope) found no association of hope with physical functioning or with symptoms and problems related to kidney disease. (9) Second, previously only hope in general had been studied (using Snyder’s hope scale), and then only among patients at stage 5D.(9) In contrast, we found quantitative differences in HR-Hope across a wide range of severity-defined categories of CKD.

Several limitations of this study also warrant mention. First, because the study was cross-sectional, we cannot infer causal relationships from the associations of HR-Hope with the stage of kidney disease, or with objective manifestations of adherence. Patients with high systolic BP might struggle with the responsibility of taking multiple antihypertensive agents and following dietary instructions, and thus they might find fluid restriction and dietary restriction to be very burdensome, which might cause them to have little or no hope. Second, we were not able to quantify adherence itself by measuring, for example, intake of salt, phosphorus, and potassium, or by counting the numbers of prescribed doses of medications that patients did and did not take. Nonetheless, it may be argued that the objective indices we used are at least as important clinically as adherence itself. Third, HR-Hope may be affected by spiritual and religious factors. This could be important in some countries, but we believe that any effects it might have had on the differences we found in this study are negligible, as all of the participants were Japanese, among whom few regularly engage in religious activities. Fourth, data on some potential predictors of dietary adherence, including detailed measures of socioeconomic status, were not collected. However, the analyses included adjustments for working status, which served as a proxy for socioeconomic status. In addition, patients’ medical costs related to dialysis treatment were almost entirely covered by Japan’s health insurance system. Thus, the associations of HR-Hope with adherence to prescribed treatment are not likely to have been confounded by socioeconomic status. Fifth, while the internal-consistency reliability of the HR-Hope scores is high, it was not possible to quantify test-retest reliability or to examine whether HR-Hope varied between dialysis days and non-dialysis days. Further examination of the HR-Hope scale’s psychometric properties is warranted.

## Conclusions

Throughout the trajectory of CKD, HR-Hope changed with disease stage. It was lowest at non-dialysis stage 5, followed by stages 4 and 5D. Health-related hope was associated with the patient-perceived burden of fluid and dietary restriction, and also with some objective manifestations of adherence to prescriptions for diet and medications. Therefore, nephrologists and dieticians should consider interventions for increasing HR-Hope to promote self-care and improve CKD patients’ adherence to those prescriptions.

## Data Availability

The datasets used and/or analyzed during the current study are available from the corresponding author on reasonable request.

**List of abbreviations**
BP: blood pressure
CI: confidence interval
CKD: chronic kidney disease
HR-Hope: health-related hope
KDQOL: Kidney Disease Quality of Life
OR: odds ratio
PS: performance status

## Declarations

### Ethics approval and consent to participate

This study was conducted in accordance with the Declaration of Helsinki. This study was approved by the ethics review boards of St. Marianna University School of Medicine (No.3209) and Fukushima Medical University (No.2417). Written consent was obtained.

### Consent for publication

Not applicable.

## Acknowledgements

The authors greatly thank the following researchers, research assistants, and medical staff members for their assistance in collecting the questionnaire-based and clinical information used in this study: Ms. Asako Tamura, Ms. Yuka Masuda, Ms. Takae Shimizu (St. Marriana University, Kawasaki-city, Kanagawa), Takayuki Nakamura, MD, Eiko Hashimoto, RN (JCHO Nihonmatsu Hospital, Nihonmatsu-city, Fukushima), Atsushi Kyan, MD, Masashi Saito, CE (Shirakawa Kosei General Hospital, Shirakawa-city, Fukushima), Ms. Lisa Shimokawa (Fukushima Medical University Hospital, Fukushima-city, Fukushima).

## Competing interests

NK, TW, YI, S. Fujimoto, M Yazawa, TS, KK, M Yanagi, HK, JG, S Fukuhara, and YS declare that they have no relevant financial interests.

## Authors’ contributions

Research idea and study design: NK, TW, YI, S Fukuhara, YS; data acquisition: NK, YI, S Fujimoto, M Yazawa, TS, KK, M Yanagi, HK; data analysis and interpretation: NK, TW, YI, JG, S Fukuhara, YS; statistical analysis: NK; supervision or mentorship: YI, JG, S Fukuhara, YS. Each author contributed important intellectual content during article drafting or revision and accepts accountability for the overall work by ensuring that questions pertaining to the accuracy or integrity of any portion of the work are appropriately investigated and resolved.

## Funding

This study was supported by JSPS KAKENHI (Grant Number: JP16H05216). The funders had no role in the study design, analysis, or interpretation of data; writing of the manuscript; or the decision to submit the manuscript for publication.

## Titles of supplementary material

Table S1. English version of the 18-item Health-Related Hope scale.

Table S2. Japanese version of the 18-item Health-Related Hope scale.

Figure S1. Conceptual framework used in regression analyses.

Table S3. The associations between HR-Hope and blood pressures.

Table S4. The associations between HR-Hope and serum phosphorus levels.

Table S5. The associations between HR-Hope and serum potassium levels

## Notes

### Competing Interest Statement

The authors have declared no competing interest.

### Author Declarations

This study was approved by the ethics review boards of St. Marianna University School of Medicine and Fukushima Medical University.

